# Novel Domain V mutation in *cfr*-positive clinical *Staphylococcus haemolyticus* isolates causing linezolid resistance in Egyptian ICU patients

**DOI:** 10.1101/2024.11.22.24317765

**Authors:** Saber Maghraby, Mohamed Ez El-Din Dawoud, Mohamed Eweis, Abdullah El-nagish, Doaa Ghaith

## Abstract

Linezolid resistance has become a focal point worldwide, particularly as linezolid stands as one of the last resort antibiotics against multiple-drug resistant bacterial strains. Despite its critical importance, the situation in Egypt remains relatively unexplored. As various linezolid resistance mechanisms have been identified in staphylococci, our investigation aims to uncover the molecular characteristics of staphylococci showing reduced susceptibility to Linezolid at Kasr-El-Eini Teaching Hospital. Thirty isolates were investigated in this study. The selected samples, identified by Vitek 2 system and confirmed by 16S rRNA, were examined by disc diffusion method. Of the 30 isolates, two were confirmed as linezolid resistant and screened for the presence of cfr gene and mutations in the Domain V of the 23S rRNA gene. Both of the studied strains, identified as *Staphylococcus haemolyticus*, SZ-2 and SZ-7 harbored two point mutations G2576T and G2602T in the Domain V of the 23S rRNA gene. A novel G2694C mutation reported for the first time was detected in strain SZ-7. The presence of cfr gene was confirmed in both isolates. Linezolid-resistant *Staphylococcus haemolyticus* had emerged in ICU patients with prior antibiotic exposure. The exact role of the novel G2694C mutation in linezolid resistance needs further investigations. The study underscores the importance of proper surveillance of *cfr*-carrying strains in the healthcare settings in Egypt.

## Introduction

*Staphylococcus haemolyticus*, a coagulase-negative staphylococci (CoNS), naturally resides in the human skin tissue microbiota with the potential to induce various illnesses, such as skin infections, bloodstream infections, and other several nosocomial infections [1]. In recent years, broad-spectrum antibacterials were widely used in clinical settings, that led to a persistent rise in drug resistance. The current surge in bacterial drug resistance is a cause for serious concern [2]. Linezolid, an oxazolidinone antibiotic, was approved for clinical use in 2000 [3]. This antibiotic is potent against multidrug-resistant Gram-positive bacteria, including methicillin-resistant *Staphylococcus aureus* (MRSA), vancomycin-resistant Enterococci (VRE), and *Streptococcus pneumoniae* [4]. Activity of linezolid results from binding to the 23S rRNA in the 50S ribosomal subunit. Thus, it acts as an inhibitor of protein synthesis by binding to the ribosomal peptidyl transferase center (PTC) affecting tRNA positioning and eventually stopping the growth of bacteria [5,6].

The prevalence of linezolid resistance is steadily on the rise due to the extensive utilization of this antibiotic. Numerous reports have documented cases of linezolid-resistant staphylococci. The most well-established mechanisms underlying the development of linezolid-resistant staphylococci involves a mutation in the 23S rRNA gene target and the acquisition of the chloramphenicol-florfenicol resistance (cfr) gene [7,8,9,10]. The 23S rRNA, a constituent of the bacterial ribosome 50S subunit, can undergo mutations during exposure to linezolid, rendering the drug ineffective and ultimately resulting in drug resistance. Additionally, gene mutation sites exhibit variability among different strains [10, 11]. Among Staphylococcus strains, G2576T gene mutation is the most prevalent [11,12,13,14]. cfr gene, first identified in plasmids of Staphylococcus isolates from German cows, encodes a methyltransferase capable of modifying 23S rRNA [15]. The cfr gene facilitates methylation at specific sites on the ribosomal large subunit, leading to resistance against linezolid [5,16].

At our Kasr-El-Eini Teaching Hospital, the escalating concern lies in staphylococcal isolates in the intensive care units, which exhibit an extraordinary predisposition to develop resistance to all available antibiotics including linezolid. Consequently, in pursuit of viable treatment alternatives, there is a pressing need to comprehend the resistance mechanism of such isolates. Henceforth, this study was undertaken to investigate the molecular characteristics of linezolid-resistant S. haemolyticus identified in clinical infections. A collection of linezolid-resistant CoNS (LRCoNS) from 30 patients were evaluated for mechanisms of linezolid resistance.

## Materials and Methods

### Sample collection, identification and antimicrobial susceptibility testing of bacterial isolates

Over two years; from March 2020 to March 2022, a total of 30 clinical isolates of staphylococci showing reduced susceptibility to Linezolid were collected from Kasr El-Eini Teaching Hospital. Samples were isolated from different specimens, including pus and blood. Disc diffusion method was used to screen isolates for linezolid resistance. Linezolid concentration used was 30 µg. The antimicrobial discs were obtained from Oxoid Ltd (Basingstoke, Hampshire, England, UK) and processed according to the manufacturer’s instructions.

The identification of isolates was performed using conventional techniques such as cultivation on mannitol salt agar [17] and deoxyribonuclease (DNase) agar [18]. The isolates were further identified using VITEK 2 system (bioMérieux, Marcy-l’Étoile, France) using the VITEK 2 GP Card according to the manufacturer’s instructions. The antibiotic susceptibility test of isolates was performed on Mueller Hinton agar using the Kirby Bauer disc diffusion method [19] as per Clinical and Laboratory Standard Institute [20] guidelines using antibiotic Linezolid 30 μg. Subsequently, identification of the selected bacterial isolates was confirmed by 16S rRNA gene [21]. All the relevant clinical data were collected for analysis, including dose and length of linezolid treatments and other results of antimicrobial susceptibility tests.

### Mechanisms of linezolid resistance among the isolates

The two most resistant *Staphylococcus haemolyticus* strains (namely SZ-2 and SZ-7) were selected and screened for point mutations in Domain V of the 23S rRNA gene and the presence of the *cfr* gene by PCR and DNA sequencing as described previously [7,8,9,10].

The genomic DNA was isolated using Quick-DNA™ Miniprep Kit from Zymo Research™ according to the manufacturer instruction manual. Amplification was performed using the following PCR conditions: 35 cycles of denaturing, annealing, and extension at 94°C (30 s), 55°C (30 s), and 72°C (1 min) respectively; and a final 10-min extension at 72°C.

To elucidate the mechanism of linezolid resistance, screening for cfr gene and point mutations in domain V of 23S rRNA gene were investigated by amplifying the loci using the PCR conditions described by Yoo et al [8]. Amplification by PCR was performed using primers listed in Table 1. Amplicons were sequenced on both strands and sequences were aligned with the corresponding sequences from linezolid-susceptible *S. haemolyticus* JCSC 1435 (GenBank accession No. AP006716). All sequenced data was uploaded to the NCBI under accession numbers shown in the data availability section.

**Table 1.**
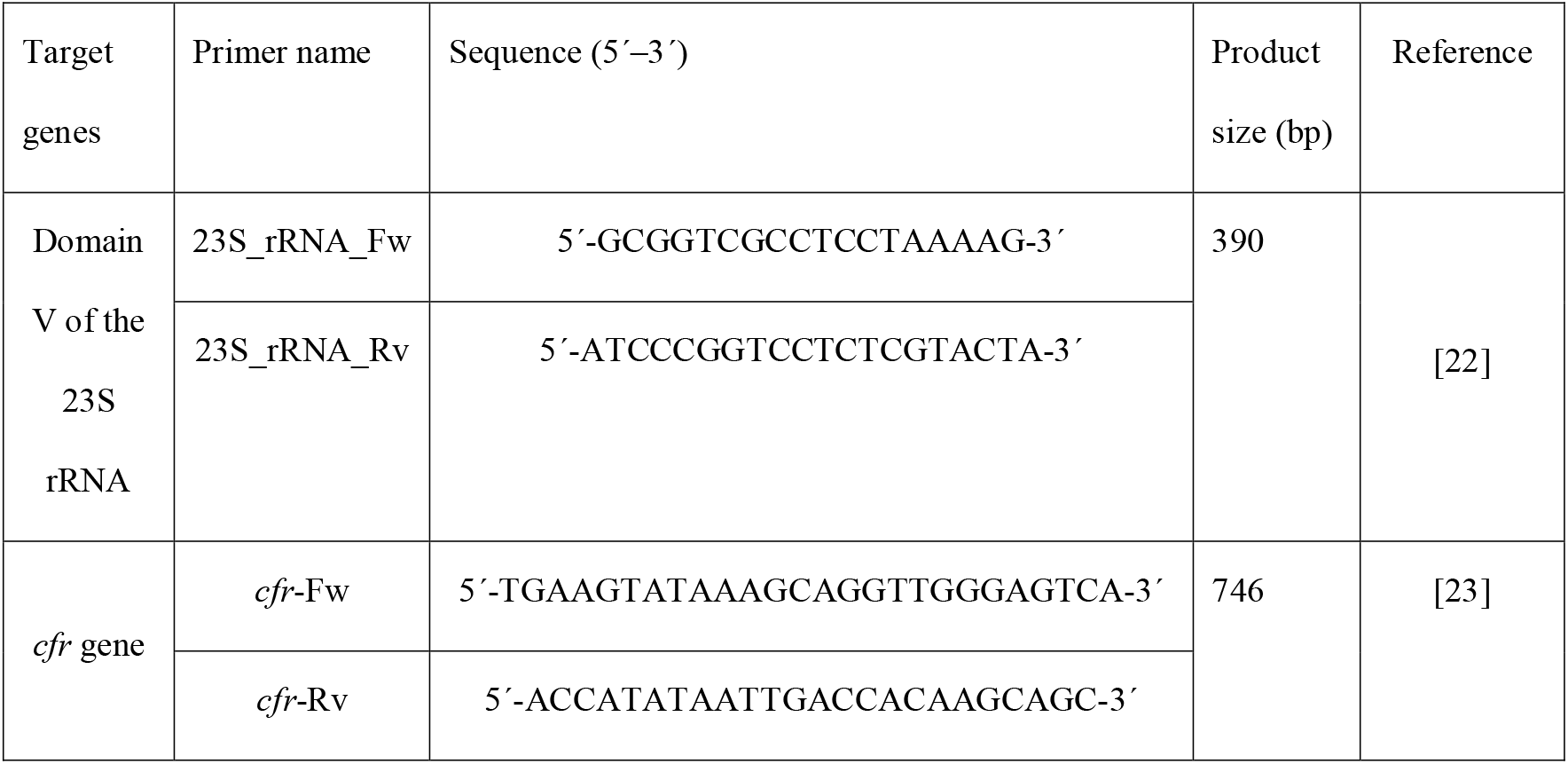
Primers are used for PCR amplification of targeted genes.

## Results and Discussion

Twenty four of the total 30 isolates were found to be totally susceptible to linezolid. Isolates SZ-2 and SZ-7, showed the highest resistance to linezolid, were selected for further investigations. The two isolates were identified as Staphylococcus based on their characteristic growth on blood agar and microscopic observations. The isolates were negative for mannitol fermentation, DNase and coagulase test, thus confirming coagulase negative Staphylococcus. The Vitek 2 system identified the isolates correctly (Table 2), with a probability of 99% and confidence level ‘excellent’. The 16S rRNA gene sequence revealed that the isolates are S. haemolyticus with 99% similarity with S. haemolyticus strain JCSC1435 in BLAST search.

**Table (2):**
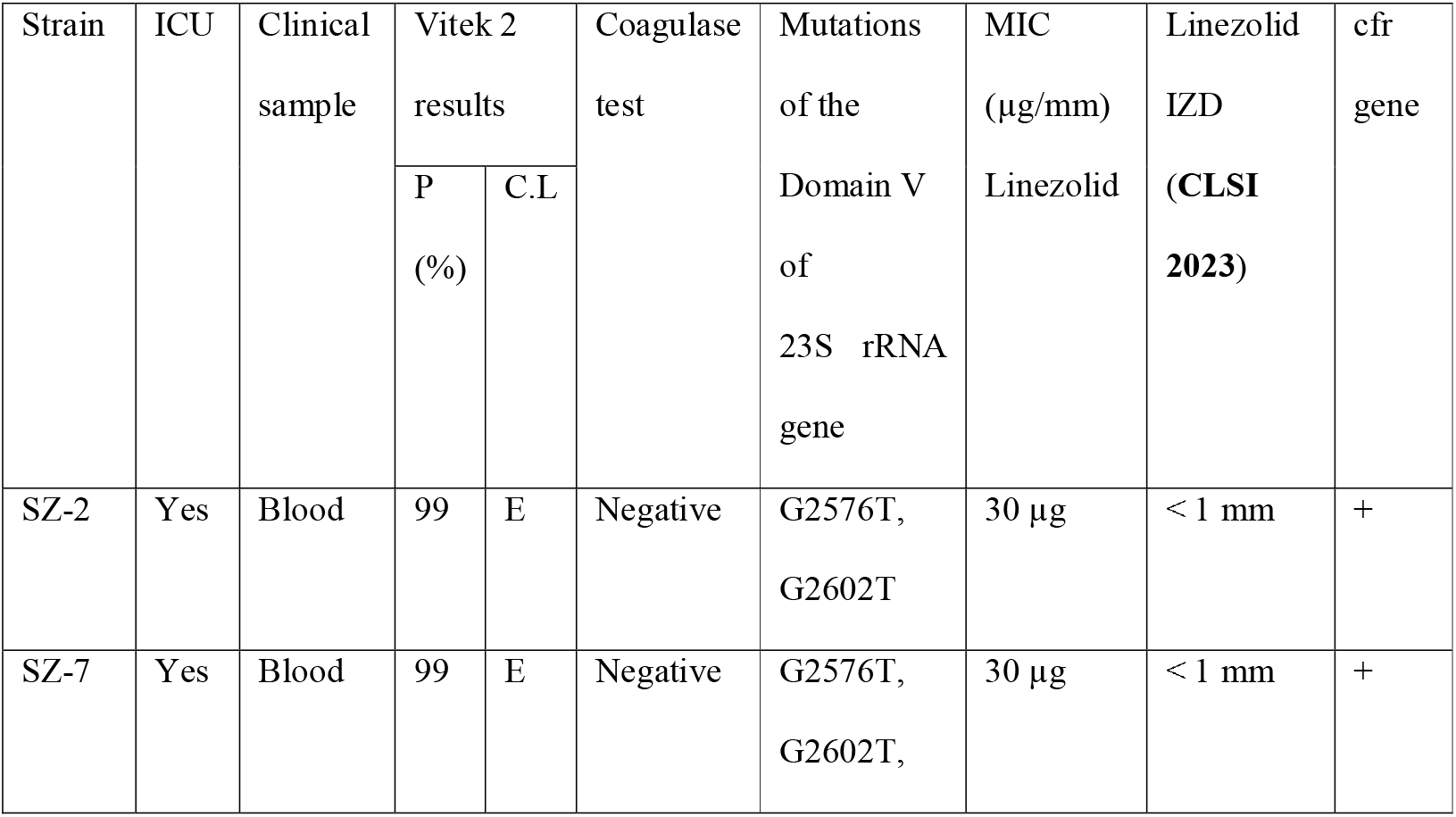
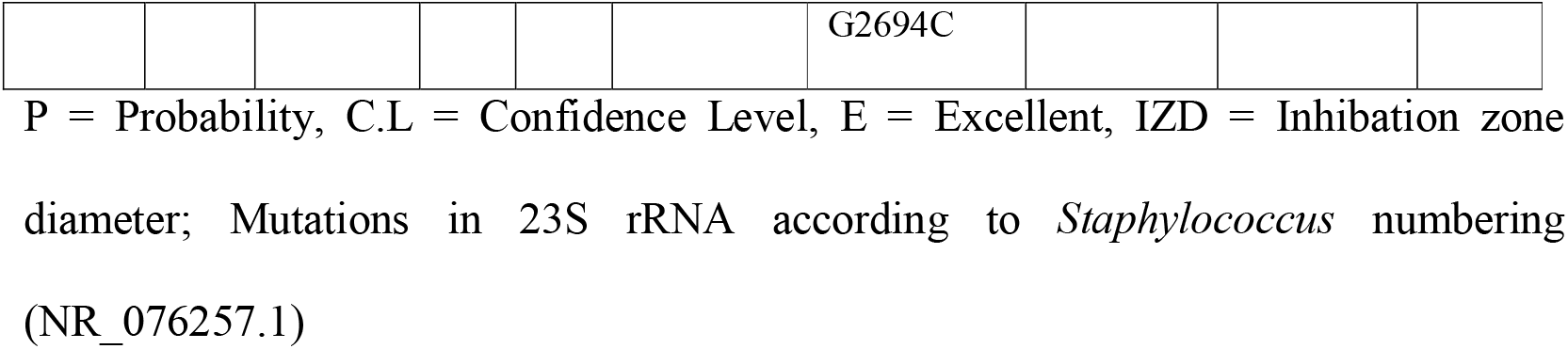
Mutations detected in the V domain of 23S rRNA gene of the resistant isolates *S. haemolyticus strain SZ-2* and SZ-7.

| Strain | ICU | Clinical sample | Vitek 2 results |  | Coagulase test | Mutations of the Domain V of 23S rRNA gene | MIC (µg/mm) Linezolid | Linezolid IZD (CLSI 2023) | cfr gene |
| --- | --- | --- | --- | --- | --- | --- | --- | --- | --- |
|  |  |  | P (%) | C.L |  |  |  |  |  |
| SZ-2 | Yes | Blood | 99 | E | Negative | G2576T, G2602T | 30 µg | < 1 mm | + |
| SZ-7 | Yes | Blood | 99 | E | Negative | G2576T, G2602T, | 30 µg | < 1 mm | + |
|  |  |  |  |  |  | G2694C |  |  |  |

P = Probability, C.L = Confidence Level, E = Excellent, IZD = Inhibition zone diameter; Mutations in 23S rRNA according to *Staphylococcus* numbering (NR\_076257.1)

Strain SZ-2, with a linezolid inhibition zone diameter (IZD) less than 1 mm (zone diameter breakpoint according to CLSI 2023 ≤ 20 mm), was found to have G2576T and G2602T mutations in the Domain V of 23S rRNA gene (Table 2). A novel G2694C mutation in addition to the same two G2576T and G2602T mutations, both in domain V of the 23S rRNA gene, were identified in *S. haemolyticus* strain *SZ-7* with a linezolid IZD less than 1 mm.

Previous studies have already reported the G2576T and G2602T substitutions in the Domain V of the 23S rRNA gene of linezolid-resistant *Staphylococcus spp*. [2,7,12,13,14,24]. Although both mutations were first reported in 2007 and in 2021, respectively [2,25], this is the first report of these mutations in *S. haemolyticus* isolated in Egypt. The novel mutation G2694C, whether it plays a role in linezolid resistance is uncertain and needs further confirmation. Nevertheless, this revelation may have the potential to contribute valuable insights into investigating the mechanism of linezolid resistance caused by mutations in the V domain of 23S rRNA gene.

*cfr* gene in both isolates was detected and confirmed by sequencing the amplicons on both strands. At Kasr-El-Eini Teaching Hospital, isolations of a *cfr*-carrying clinical *S. haemolyticus* isolates from the ICU units is extremely alerting to us. Our greatest concern is the presence of the *cfr* gene in both isolates, as its minimal fitness cost allows cells to retain it even in the absence of selection pressure from the antibiotic. Typically, the cfr gene is situated in an unstable genetic environment, either on the chromosome or on multidrug-resistant plasmids [26]. This precarious positioning would facilitate the easy spread of cfr into susceptible populations and other pathogenic bacteria. Moreover, *cfr*-mediated resistance confers resistance to a wide-spectrum of antibiotics not only linezolid and thus poses a significant challenge to therapeutic options.

Although linezolid resistance due to mutations in Domain V of the 23S rRNA and presence of cfr gene were reported in several staphylococcal species form many countries around the world such as Spain [27], the USA [28], Brazil [29], Mexico [30], Japan [31] and Korea [32], this is the first report of these mutations in S. haemolyticus isolated in Egypt. This first emergence of LRSH in intensive care units in our hospital during a short period is very worrying, requiring serious measures and proper surveillance in the healthcare settings to prevent the intrahospital dissemination of resistant strains. The novel G2694C mutation in Domain V of the 23S rRNA, whether it plays a role in linezolid resistance is uncertain and needs further confirmation. Nevertheless, this revelation may have the potential to contribute valuable insights into investigating the mechanism of linezolid resistance caused by mutations in Domain V of the 23S rRNA. The study emphasizes the need for proper surveillance of cfr-carrying strains in the healthcare settings in Egypt.

## Data availability

All sequenced data was uploaded to the NCBI under accession numbers PP525131-PP525132 for *cfr* genes and PP506638-PP506639 for 23S ribosomal RNA gene of *S. haemolyticus* strain SZ-2 and SZ-7 respectively.

## Acknowledgements

Not applicable.

## Ethical approval

Ethical approval was granted by the Review Board of the Clinical and Chemical Pathology Department, Faculty of Medicine, Cairo University, Egypt prior to the beginning of the study with a waiver of informed consents.

